# Deep Learning Approach to Measure Alveolar Bone Loss After COVID-19

**DOI:** 10.1101/2023.11.20.23298788

**Authors:** Sang Won Lee, Kateryna Huz, Kayla Gorelick, Thomas Bina, Satoko Matsumura, Noah Yin, Nicholas Zhang, Yvonne Naa Ardua Anang, Jackie Li, Helena I. Servin-DeMarrais, Donald J McMahon, Michael T. Yin, Sunil Wadhwa, Helen H. Lu

## Abstract

Severity of periodontal disease may be determined by measurement of alveolar crestal height (ACH) on dental bitewing radiographs; however, the prevailing method of assessment is through visualization which is time consuming and not a direct measure. The primary objective of this manuscript is to create and validate a deep learning technique for precise evaluation of alveolar bone loss in bitewing radiographs. Additionally, surveys were conducted with dental professionals to determine accuracy of visualized measures of ACH for severe periodontal disease versus the deep learning program and to determine the acceptability of utility of the program among diverse dental professionals. Lastly, the deep learning program was utilized in research to evaluate the role of COVID on periodontal disease through longitudinal measures of bitewing radiograph ACH from patients during the: "pre-pandemic" (Feb 2017 - Feb 2020) and "post-pandemic" (Feb 2020 - Feb 2023) periods. The pre-pandemic group had a mean percentage loss of ACH of -1.74 + 16.5%, representing a gain in alveolar bone. In contrast, the post-pandemic group had a gain in ACH of 2.46 + 14.6%, representing a loss in alveolar bone. There remained a trend for greater annualized percent change in ACH in the post-pandemic vs pre-pandemic group (1.33 + 11.9% vs -0.94 + 12.5%, p=0.07), after accounting for differences in duration between xrays. Overall, this study demonstrates the successful training and validation of a deep learning program for ACH measurement as well as its utility and acceptability among dental professionals for clinical and research.

## I. Introduction

Various medical domains have widely employed deep learning (DL) models, encompassing tasks such as the identification of anatomical structures and the detection of pathological findings on radiographic images [1–3]. In dentistry, advanced DL models present an opportunity for an objective and dependable means of diagnosis clinically significant periodontal disease and tracking progression of periodontal disease. Alveolar crestal height (ACH), defined as the distance in mm between the cemento-enamel junction (CEJ) and the alveolar crest (AC) visualized on dental xrays, is an objective measure of alveolar bone loss associated with periodontal disease. Based upon the American Academy of Periodontology update to the 1999 Classification of Periodontal Diseases and Conditions, severe (advanced) periodontal disease can be classified by ACH of >5mm [4]. The prevailing method for measuring ACH on xrays involves visually analyzing intra-oral radiographs, which is prone to errors [3]. Numerous research efforts have employed deep learning models to assess alveolar bone levels using panoramic radiographs [3, 5–7]. Despite this, a standardized approach for measuring alveolar bone levels has yet to be universally agreed upon. In our study, we detail the development and training of deep learning algorithms, leveraging training, validation, and test datasets that have been carefully curated by an oral radiologist. The primary objective of this study was to 1) introduce a new DL-based program for automatic calculation of the ACH (Fig. 1), 2) conduct a survey among dental professionals regarding their usage of this app, and 3) demonstrate the utility of the program as a research tool through a case study of longitudinal change in ACH among dental patients before and after the COVID pandemic.

**Figure 1.**
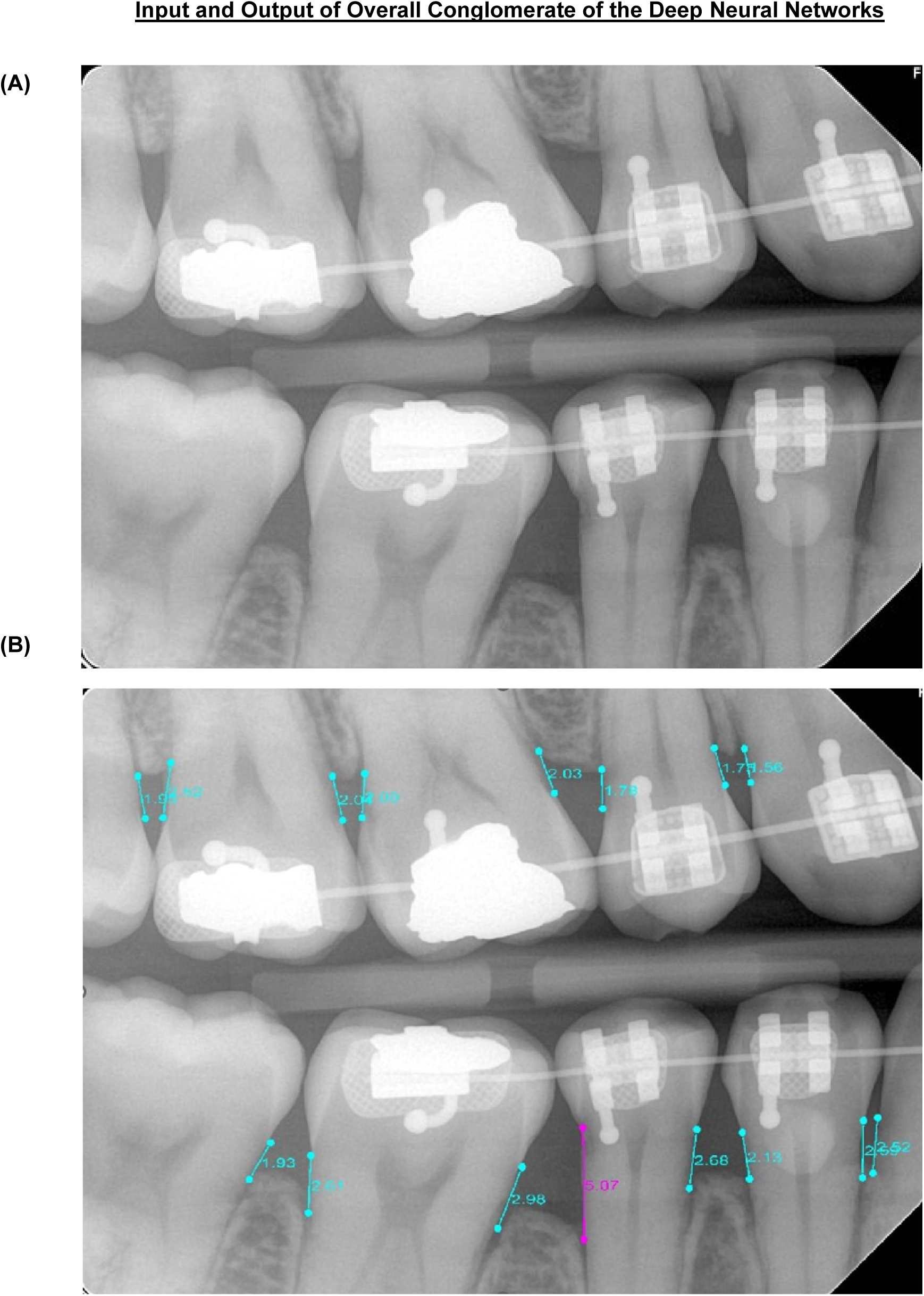
Input and Output of Overall Conglomerate of the Deep Neural Networks. Input bitewing radiographs are fed into sequential deep neural networks to produce output images overlaid with the calculated ACH length measurements. ACH greater than 5mm is highlighted in magenta.

COVID originated as a pneumonia outbreak in the province of Wuhan, China in December 2019. The virus was declared a pandemic on May 11, 2020 by the World Health Organization (WHO) [8]. During the year of 2020, the New York City hospital system was overflowing with patients who were diagnosed with COVID. The most common routes of transmission of COVID are direct transmission such as cough, sneeze and droplet inhalation. However, the virus may also be transmitted through contact with oral, nasal and eye mucous membranes [9]. SARS-COV-2 is a single stranded RNA virus which has a spike protein (S-protein) which allows it to adhere and invade the host cells [10]. The S-protein binds to angiotensin-converting enzyme 2 (ACE-2) which is seen in the lungs, kidneys and myocardial cells [11]. In addition to the lungs, kidneys and myocardial cells, ACE-2 is also found intraorally on the salivary glands and the tongue, making the oral cavity one of the first entry points [12]. Periodontitis a common cause of dental alveolar bone loss. Periodontal disease is a chronic inflammatory condition which is caused by several microorganisms such as F. nucleatum, P. gingivalis and P. Intermedia [13]. There have been many studies completed on the relationship of periodontal disease and systemic conditions. Periodontal disease and COVID share similar risk factors and comorbidities which may increase the severity of the outcomes [14].

In a previous study examining the relationship of periodontal disease and COVID, we found that patients who were diagnosed with COVID by a positive PCR test had higher ACH than a control group with a positive COVID test, and greater change in ACH suggestive of greater alveolar bone loss in the 2 years preceding their COVID positive test [15]. However, these findings may reflect a recruitment bias, since patients receiving COVID diagnoses early in the pandemic were also more likely to have co-morbidities such as hypertension, cardiovascular disease and diabetes [16]. The patient population with a higher risk of complications secondary to COVID may have also experienced dental alveolar bone loss due to other pre-existing periodontal comorbidities [17]. In this follow up study, we will perform longitudinal analyses of ACH from dental xrays of patients from the same clinic as our prior study, using the DL model, and compare change in ACH in pre-pandemic” group with patient data spanning from February 1st, 2017 to February 1st, 2020 and the second “post-pandemic” group contained patient data spanning from February 1st, 2020 to February 1st, 2023. Our hypothesis was that there would be greater alveolar bone loss in the post-pandemic vs. the pre-pandemic group.

## II. Materials and Methods

### 1. Artificial Intelligence-Deep Learning

In this study, a conglomerate of five deep neural networks was used to automate the ACH levels in bitewing radiographs. Initially, two object detection networks were employed to locate the coordinates ABCL and CEJ in the input radiographs (Fig. 2B). Subsequently, a semantic segmentation neural network was used to determine the pixel positions of ABCL (Fig. 2C), following which a best-fit line was drawn between the ABCL of the maxillary and mandibular arches to partition the upper and lower ABCL and CEJ (Fig. 2D). Polynomial curves were then fitted to the ABCL and CEJ points of both arches (Fig. 2E). Further, two semantic segmentation neural networks were utilized to identify the pixel locations of teeth in the radiographs (Fig. 2F). The tooth outlines from the previous tooth segmentation were then used to draw ACH = CEJ-ABCL if the outlines crossed both polynomial lines (Fig. 2G). Finally, the ACH levels were superimposed on the original input bitewing radiographs and provided as outputs (Fig. 2H). Examples of inputs and outputs of the conglomerate networks are shown in Figure 1.

**Figure 2.**
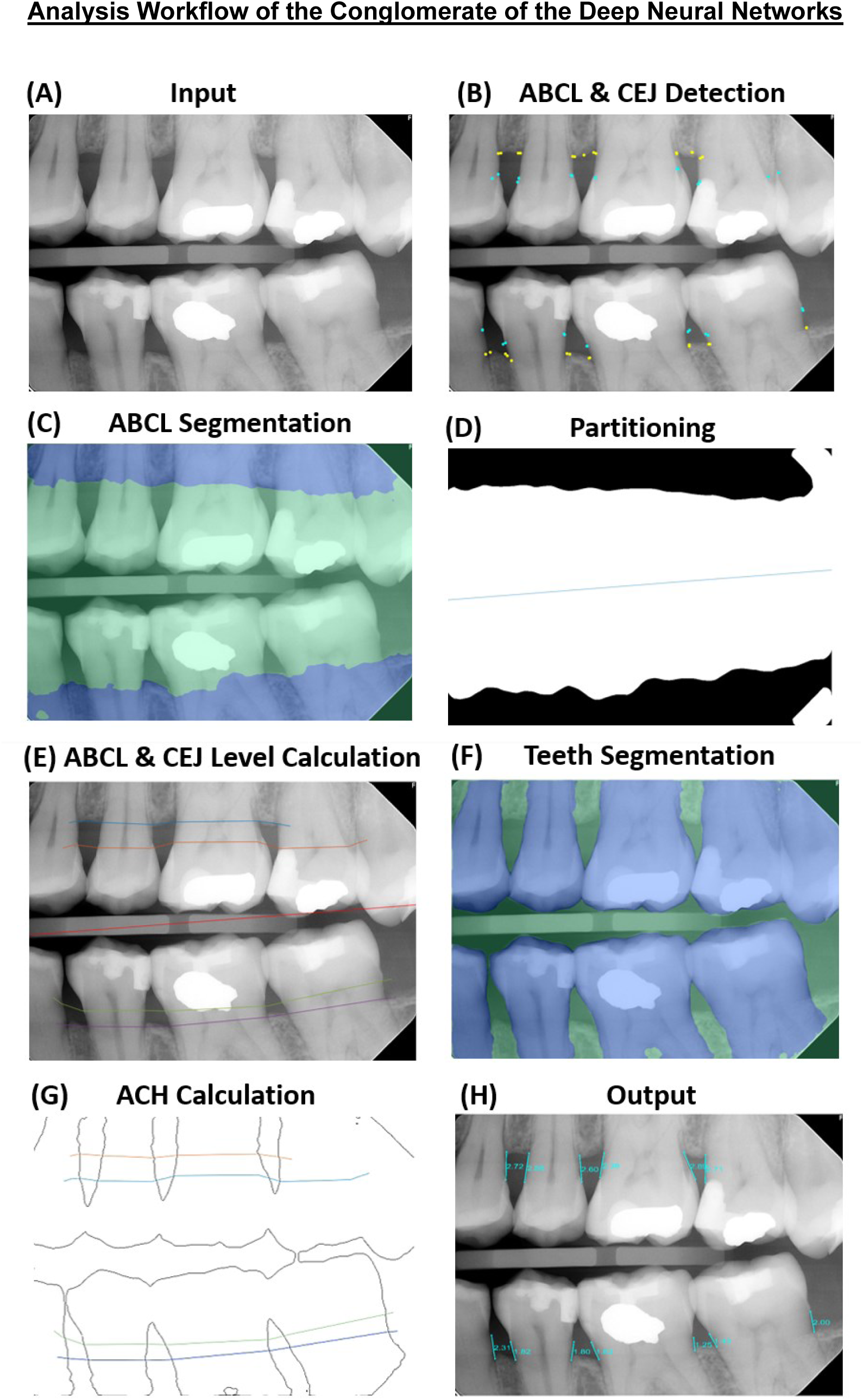
Analysis Workflow of the Conglomerate of the Deep Neural Networks. **(A)** Input bitewing radiograph. **(B)** Two object detection networks located the coordinates of ABCL and CEJs in the input radiographs. **(C)** A semantic segmentation neural network determined the pixel positions of ABCL. **(D)** A best-fit line was drawn between the ABCLs of the maxillary and mandibular arches to partition the upper and lower ABCLs and CEJs. **(E)** Polynomial curves were then fitted to the ABCL and CEJ points of both arches. **(F)** Two semantic segmentation neural networks identified the pixel locations of teeth in the radiographs. **(G)** The tooth outlines were then used to draw ACHs. ACH was determined by tooth outline intersecting both ABCL and CEJ lines. **(H)** The ACH levels superimposed on the original input bitewing radiographs and provided as outputs.

#### 1.1 Object detection deep neural networks

Two object detection neural networks were coded in MATLAB. For each of the two networks, an Inception-ResNet-v2 network was transformed into a Faster-R-CNN network by adding a region proposal network, ROI max pooling layer, classification, and regression layers to support object detection [18, 19]. The key elements of Faster R-CNN are the region proposal network (RPN) and the region-based convolutional neural network (CNN). The RPN generates a collection of potential object regions, also known as regions of interest (RoIs), that are probable to contain an object. Subsequently, the region-based CNN analyzes each RoI to classify it into distinct object categories and optimize its bounding box coordinates. The fully convolutional RPN is fed an image as input, and it generates a list of rectangular object proposals, along with their corresponding objectness scores. By sweeping a sliding window over the image, the network produces a group of k-anchor boxes (bounding boxes of various sizes and aspect ratios that are predetermined). For each anchor box, the RPN computes the likelihood of it containing an object, along with the displacement required to align with the object’s ground-truth bounding box. These predictions are used to form a group of RoIs, which are then forwarded to the region-based CNN. The rectified linear unit (ReLu) at the 625th layer was used as a feature extraction layer, and a region of interest pooling layer with an output size of [17 17] was inserted after this feature layer. The object detection neural networks were trained, validated, and tested on 550 bitewing radiograph images that were annotated by an Oral Radiologist from Columbia University Dental College. The images were randomly partitioned into training, validation, and testing sets in a 7:2:1 ratio for each of the two neural networks. The ABCLs and CEJs were enclosed by 60x60 pixel boxes, and the coordinates of these boxes were used in the training, validation, and testing sets. All images were resized to [869 1200 3] because the first object detection neural network layer was modified to accept this specific dimension. One neural network was trained for 20 epochs with mini-batch sizes of 2 and an initial learning rate of 1e-3. Another neural network was trained for 10 epochs with equal parameters. The object detection models were trained to predict optimal boxes centered on the ABCL and CEJ of teeth from an image, the centers of which were interpreted as the location of the ABCL and CEJ and subsequently noted by small circles with colors corresponding to their label in the output image. Due to the random partitioning of the training, validation, and testing sets, both networks were utilized for detecting ABCLs and CEJs as they demonstrated different levels of performance. Specifically, one of the networks achieved an average precision of 0.60 for all recalls in detecting ABCLs and an average precision of 0.65 for all recalls in detecting CEJs. On the other hand, the other network achieved an average precision of 0.60 for all recalls in detecting ABCLs and an average precision of 0.72 for all recalls in detecting CEJs. Both networks were employed to address the limitations of each other in detecting the ABCLs and CEJs in certain regions. Training the object detection models were trained on Nvidia Titan X graphic processing unit (GPU).

#### 1.2 Semantic Segmentation deep neural networks

Three semantic segmentation deep neural networks were coded in MATLAB. DeepLab v3+ convolutional neural networks for semantic image segmentation were created using Inception-ResNet-v2 as a base network [18, 20]. The input layer was coded to take input bitewing radiographs with a size of [1000 1000 3] for the network that segments ABCL and [500 500 3] for the networks that segment teeth. One deep neural network was tasked with creating semantic masks of ABCL; thus, the network was modified to match the number of output label classes: ABCL and non-ABCL. Two deep neural networks were tasked with creating semantic masks for teeth; thus, the networks were modified to match the number of output label classes: Teeth and non-teeth. The random partitioning of the training, validation, and testing sets resulted in the utilization of both networks for segmenting teeth, as they exhibited varying levels of performance. One of the networks achieved a global accuracy of 0.9567, while the other network achieved a global accuracy of 0.9562. Both networks were employed to address the limitations of each other in detecting the segmentation of teeth in certain regions.

The DeepLab v3+ networks were designed using an encoder-decoder architecture, which involves encoding the input bitewing radiograph image into a lower-dimensional representation and then decoding it to generate the output segmentation mask. This architecture was preferred for the semantic segmentation models because it can handle large bitewing radiograph sizes while still preserving fine-grained details. In addition to the encoder-decoder architecture, dilated convolutions were used to increase the receptive field of image the network, enabling it to capture contextual information from a larger area in the input bitewing radiographs. Skip connections, which connect the output of the encoder to the decoder without downsampling to help preserve high-resolution features in the overall output. Overall, the combination of these techniques and the Inception-ResNet-v2 base network resulted in a powerful semantic segmentation network capable of accurately segmenting bitewing radiographs.

To train, validate, and test the networks, a total of 500 bitewing radiographs were used. Label images corresponding to the bitewing radiographs were manually created with masks identifying ABCLs at pixel level for the semantic segmentation neural network that was tasked with segmenting ABCL. Label images corresponding to the bitewing radiographs were also manually created with masks identifying teeth in pixel level for the semantic segmentation neural networks that were tasked with segmenting teeth. DeepLab v3+ was trained using 60% of the bitewing radiographs from the dataset. The rest of the bitewing radiographs were split evenly into 20% each for validation and testing, respectively. To balance the classes for the training images, median frequency class weights were used by counting pixel labels, and the classes were weighed accordingly. Stochastic gradient descent with momentum was used for training. The neural network for ABCL segmentation was trained for 146 epochs with mini-batch sizes of 4 and an initial learning rate of 1e-3. The neural networks for teeth segmentation were trained for 150 epochs with mini-batch sizes of 4 and an initial learning rate of 1e-3. The segmentation models were trained using Nvidia Titan RTX GPU.

### 2. Survey

This study was approved by the Columbia University International Review Board (IRB), reference number AAAT2272. To evaluate the accuracy and efficiency of manual xray examination done by humans and juxtapose these findings with the outcome of our software, a twenty question survey was created. This survey was sent to members of Columbia Dental School’s alumni community that included orthodontists, general dentists, dental students. Periodontists, and Endodontists. The survey was designed to ask questions that gave a general overview of the level of expertise of the participant while also testing their knowledge and efficiency at identifying alveolar bone loss using ACH measurement through test xray images. It also presented each participant with our computer program’s reading of the xray images and asked them to judge its evaluation of the xrays. The accuracy results of the survey were then compared to the AI accuracy, with both being compared to the gold standard: the Columbia Dental School Oral Radiologist.

To evaluate the expertise level of each participate, the survey was designed to inquire about the dentistry practice area of the participant, their professional setting, their duration of practice, and whether or not they use software to assist in the reading of xrays in their practice. The survey was modeled to then present each participant with the same three test xrays and ask them to read and evaluate the severity of bone loss by measuring the ACH for both the medial and distal side of each tooth, defining a measurement of <5mm as not severe and one of ≥5mm as severe. After each test xray, the participant was asked to list the number of minutes it took them to read the xray. The survey also introduced them to our AI computer algorithm that read the images itself, asking them to evaluate the accuracy of the program and any major differences between their readings and the computer’s.

### 3. COVID: Study population

This study was approved by the International Review Board (IRB), reference number AAAT2272. Dental records, medical records and radiographs are all accessed through the same electronic records platform (EPIC) at Columbia Irving Medical Center (CUIMC). This study utilized a database of patients who had dental procedures at CUIMC between 2/1/17 and 2/1/23, who received at least one PCR test for SARS-CoV-2 at CUIMC.

We created two groups of patients: one “pre-pandemic” group with patient data spanning from February 1st, 2017 to February 1st, 2020, and “post-pandemic” group with patient data spanning from February 1st, 2020 to February 1st, 2023. To be included in either the pre-pandemic or post-pandemic group, a patient from the database must have had bitewing images from two dates within their respective cohort time period. In order to increase the accuracy of the comparison of alveolar bone loss over time, the bitewing images across the two dates were taken from the same side of the patient’s mouth. In total, 231 patients met criteria for the pre-pandemic group and 163 patients met criteria for the pre-pandemic group, and were included in a case-control, cross sectional analysis. In addition, 71 of patients in the pre-pandemic group also had a pair of bitewing xrays obtained in the post-pandemic period; therefore, data from these patients were utilized in a separate longitudinal analysis.

### 4. COVID: Medical and Dental chart Review

Medical and Dental data was collected manually from EPIC for all 394 patients. Age, Sex, Race, ethnicity, diabetes diagnosis, HIV diagnosis and smoking status were recorded from patients charts in EPIC. The smoking status was recorded as ever use.

Alveolar Crestal Height (ACH) was measured utilizing bitewing radiographs. The bitewing radiographs for each patient were collected from Medcore Imaging MIPACS Dental Enterprise Viewer Software. Once the images were collected and sorted into the pre- and post-pandemic groups, the radiographs were analyzed using an artificial intelligence program that utilized landmarks on each tooth in the bitewing image to measure alveolar crest height (ACH) in multiple sites across the images. ACH was measured as the distance between the CEJs and the alveolar crest (AC) on the mesial and distal sides of the all the teeth present in a given bitewing xray. ACH measurements from each visualized tooth were then averaged to create a mean ACH level for each bitewing xray. Mean ACH levels from the same patient were subtracted across the two collection dates to find the mean ACH loss for that particular patient over the two collection dates. The mean ACH loss across the pre-pandemic and post-pandemic group were then compared in order to analyze the difference in mean bone loss between groups, investigating the potential connection between the COVID pandemic with an increased rate of bone loss over time.

Additional details regarding object detection and semantic segmentation deep neural networks can be found in Supplementary Materials.

### 5. COVID:Statistical approach

Overall differences in demographics (age, race, ethnicity) and comorbidities associated with periodontal disease (smoking ever, diabetes) between COVID+ and pre-pandemic controls were evaluated with Chi-square or Fisher’s Exact Test for categorical variables and T-tests or Wilcoxon Sign Rank tests for continuous variables. A missing value for race, ethnicity, smoking and diabetes was represented in the reference coding and analysis. Results were reported for the non-missing coding against the reference group: white, non-Hispanic, non-diabetes, non-smoker, respectively. A total of 394 unique individuals had pairs of bitewing xrays available for calculation of ACH.

The following outcomes were calculated. Mean ACH (mm) was calculated by taking the sum of all ACH for each tooth visualized on the bitewing divided by the number of evaluable measurement sites. Change in ACH was calculated by subtracting mean ACH of first xray from second xray. A positive change in ACH reflects an increase in ACH over time, which is equivalent to a loss of bone over time. A negative change in ACH reflects a decrease in ACH over time, which is equivalent to a gain in bone over time. Percent change in ACH was calculated by dividing change in ACH by the mean ACH from the first bitewing. Annualized percent change in ACH was calculated by dividing the percent change in ACH by years between first and second bitewing.

Cross-sectional differences in annualized percent change in ACH between post- and pre-pandemic controls were further examined for the influence of participant sex and age, ethnicity and age, and race and age using separate general linear models for the main effects and the interaction: age entered as a continuous variable or coded in decades and multiple comparisons adjusted by the method of Scheffé. Longitudinal analysis of the difference in post- and pre-pandemic xrays used within-subject paired T-tests. The influence of sex, age, race and ethnicity on the longitudinal difference in ACH loss used multiple regression with variable coding for main and interaction effects as describe for the general linear models, above. No missing data were imputed and adjustment for multiple comparisons was restricted to within model comparisons between model-estimated least squares means. All data management and statistical analyses used SAS (SAS Institute, Cary, NC).

## III. Results

### 1. Computer Application: Deep Learning Results

In this study, two distinct semantic segmentation deep learning algorithms were utilized for specific tasks. The first algorithm aimed to segment teeth, achieving a global test set accuracy of 0.9567. The second algorithm focused on segmenting alveolar bone and attained a commendable global test set accuracy of 0.9281. For object detection, two separate algorithms were employed. The highest average precision of all recalls achieved by an algorithm, designed for detecting cementoenamel junctions, was 0.72. As for the second algorithm, intended for detecting alveolar bone crestal levels (ABCLs), it achieved the highest average precision of all recalls at 0.65.

### 2. Survey Results

#### 2.1 Deep learning vs. Dental Professionals

In this study we compared the measurement of ACH on all visible teeth in three different bitewing xrays by 56 different dental professionals against the deep learning application’s performance. An example of deep learning analysis on an input bitewing xray is shown in Figure 1. The evaluation focused on the DL-application’s ability and dental professionals’ ability to classify severe periodontal disease (ACH >5mm) vs. non-severe (<=5mm) ACHs using 35 calculatable ACHs present in three xray images of the tooth. The application achieved 94% accuracy in its classifications, surpassing the performance of the dental professionals, who achieved a 68% accuracy rate. Further analysis of individual xray images revealed the following results (Fig 3): In the first xray image, there were 16 calculatable ACHs. Dental professionals correctly classified 87% of teeth with severe periodontal disease (SD=16%), while the deep learning application demonstrated perfect accuracy with 100% correct classifications. The time that the participants took to analyze the xray had a mean of 105.3 seconds (SD=68.7) and a median of 60 seconds. In the second xray image, there were 6 calculatable ACHs. Dental professionals accurately identified only 35% of teeth with severe periodontal disease (SD=29%), whereas the deep learning application achieved an 83% accuracy rate. The time that the participants took to analyze the xray had a mean of 71.2 seconds (SD=42.6) and a median of 60 seconds. In the third xray image, there were 13 calculatable ACHs. Dental professionals correctly classified 82% of teeth with severe periodontal disease (SD=13%), and the deep learning application displayed performance with a 100% accuracy rate. The time that the participants took to analyze the xray had a mean of 80.6 seconds (SD=52) and a median of 60 seconds. All images took less than 10 seconds for the deep learning application.

**Figure 3.**
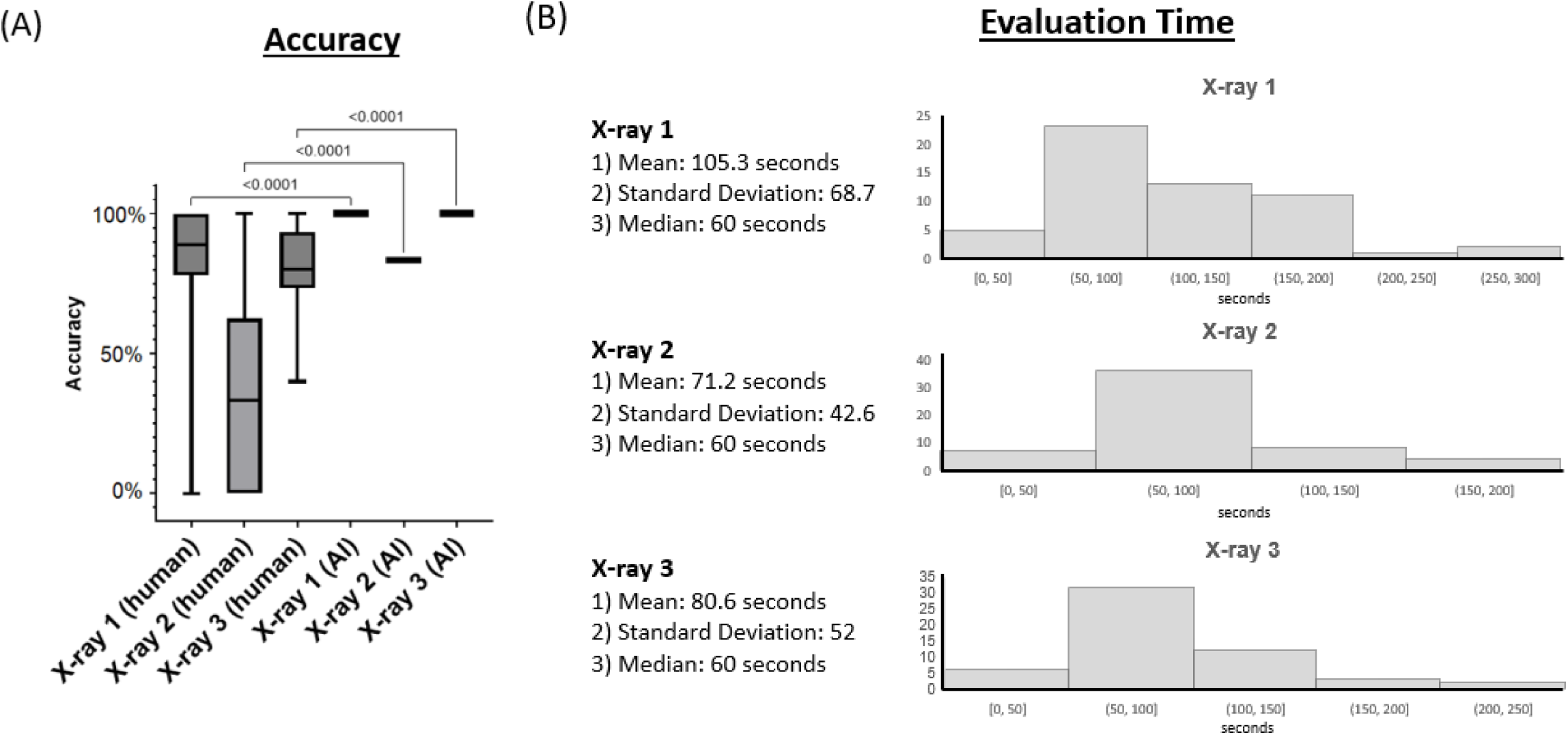
DL vs. Dental Professionals. **(A)** DL-application outputs and dental professionals’ (n=56) outputs were compared against the gold standard. **(B)** Mean, standard deviation, median, and histograms of time taken by the participants in analyzing the xrays.

#### 2.2 Acceptability of deep learning software from dental professionals

In total, the dental survey was fully completed by 64 participants. (Fig 4 and 5) Of the responses, 34% came from orthodontists, 30% came from general dentists, and 16% came from dental students. The remaining 20% of responses came from other dental professions including Periodontists and Endodontists. More than half the participants came from an academic setting--totaling at 52%--while 33% came from private practices, 8% came from group practices, and 7% came from another dentistry work setting. Automated or deep learning software was only used by 21% of the participants to assist in their reading of xrays, and a majority of 57% stated that they only approximate when measuring bone levels--compared to the 9% that measure with a ruler. The results of the survey indicated that 51% of participants agreed with the AI’s ACH measurements, with an extra 33% strongly agreeing and no participants disagreeing. Furthermore, 56% of participants agreed that AI would be helpful in their professional setting.

**Figure 4.**
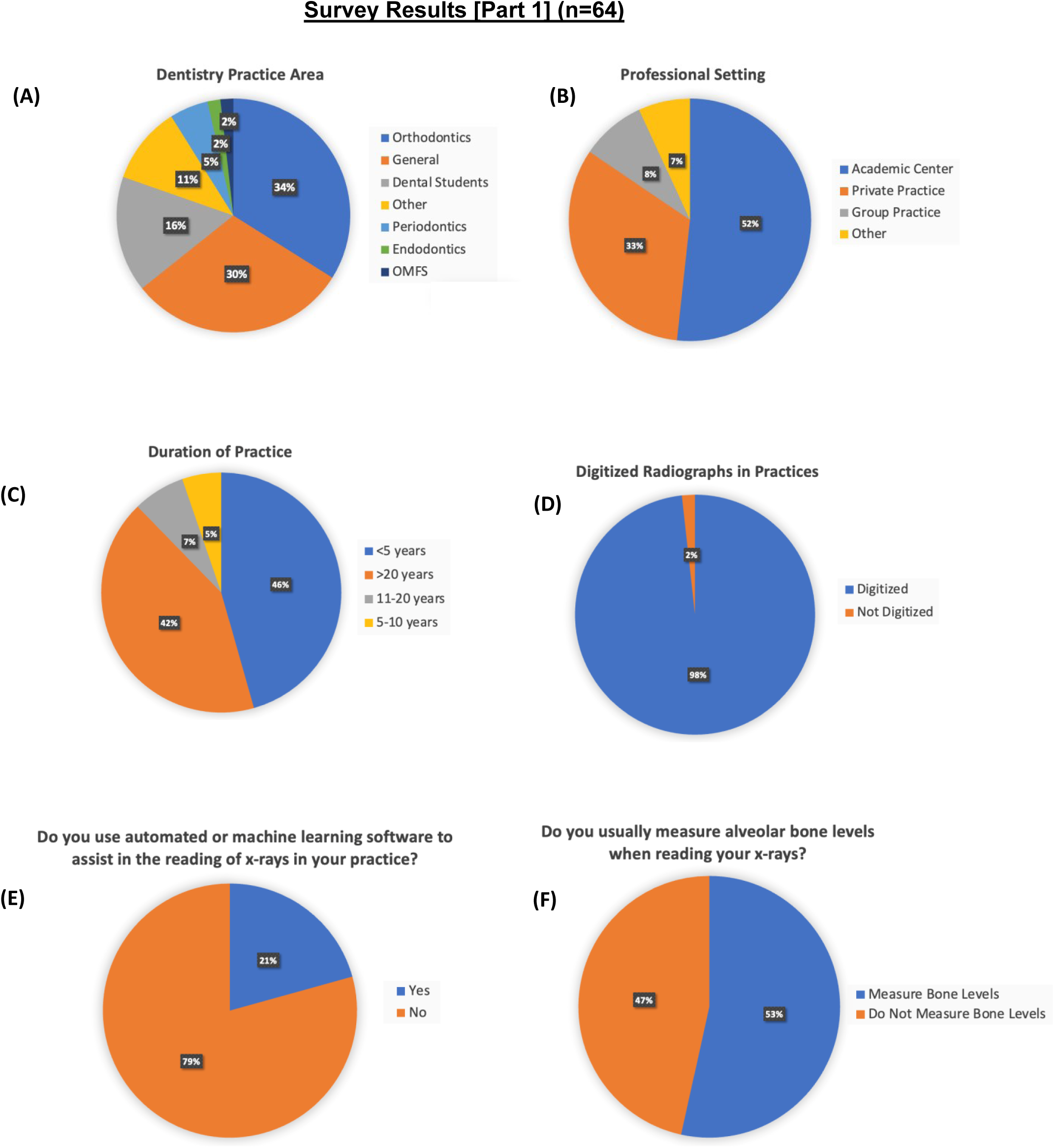
Survey Results (Part 1). 64 participants from Columbia Dental School alumni were surveyed regarding their dentistry background and their methods for reading xrays.

**Figure 5.**
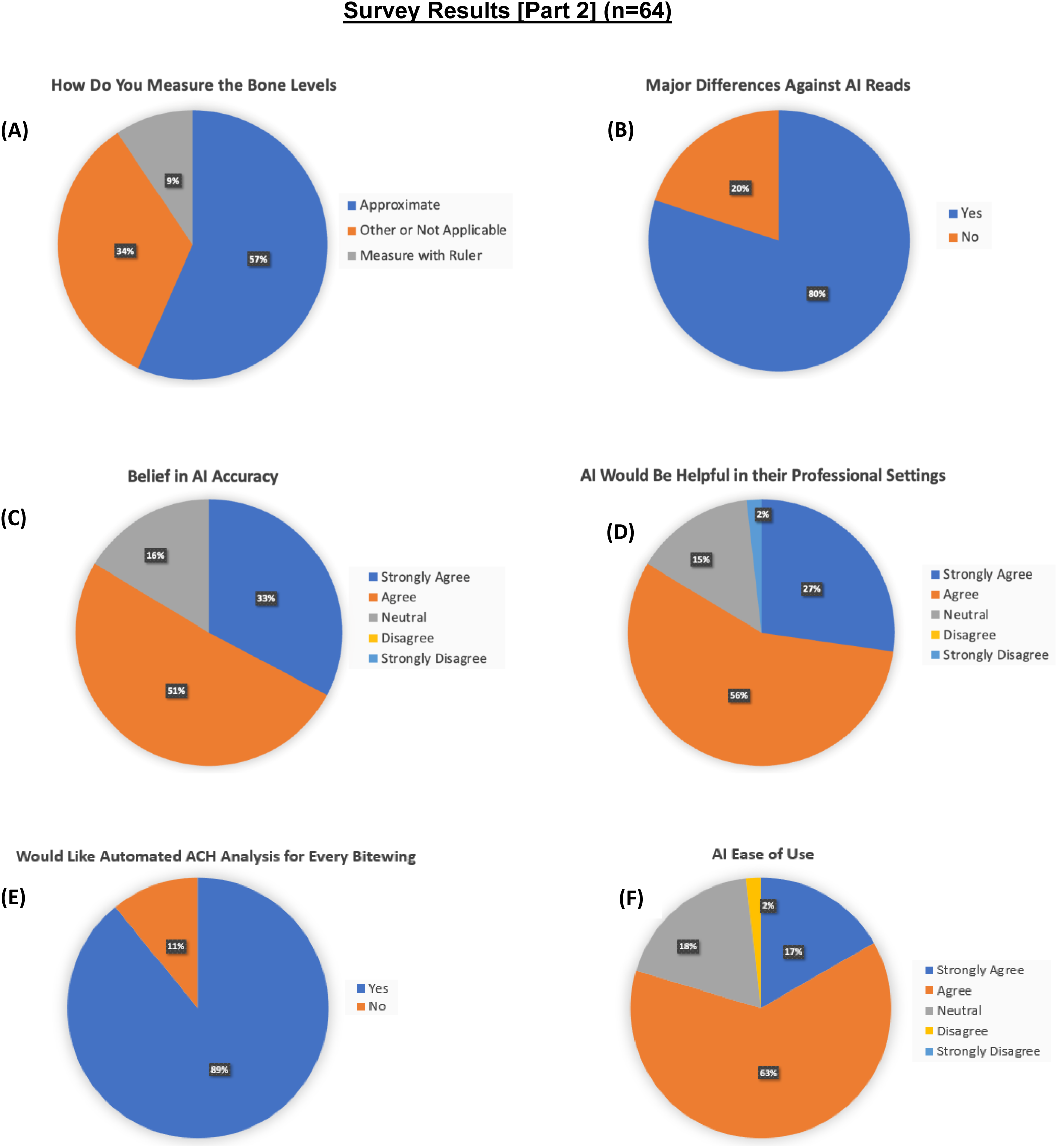
Survey Results (Part 2). 64 participants from Columbia Dental School alumni were surveyed regarding their methods for measuring bone levels and their opinion on the use of AI to assist them.

In our study sample, age, sex, race/ethnicity, history of smoking (ever), and history of diabetes and HIV were similar in the pre-pandemic and post-pandemic groups (Table 1). The pre- and post-pandemic groups also did not differ with regards to the mean ACH or mean number evaluable sites on bitewings at baseline (Table 1).

**Table 1.** Demographics, risk factors, baseline ACH and change in ACH in pre-pandemic and post-pandemic groups.

|  | **Pre-pandemic**<br>**(N=231)** | **Post-pandemic**<br>**(N=163)** | P value |
| --- | --- | --- | --- |
| Age | 50.1 ± 19.4 | 49.1 ± 19.5 | 0.61 |
| Sex |  |  | 0.58 |
| Male | 66 (28.6%) | 54 (33.1%) |  |
| Female | 164 (70.9%) | 108 (66.3%) |  |
| Unknown | 1 (0.4%) | 1 (0.6%) |  |
| Race |  |  | 0.75 |
| White | 63 (27.3 %) | 52 (31.9%) |  |
| Black | 36 (15.6%) | 27 (16.6 %) |  |
| Asian | 7 (3.0 %) | 3 (1.8 %) |  |
| Other | 125 (54.1%) | 81 (49.7%) |  |
| Hispanic ethnicity | 148 (64.0 %) | 95 (58.0%) | 0.32 |
| Smoking (ever) | 59 (25.5%) | 33 (20.2%) | 0.33 |
| Diabetes | 33 (14.3 %) | 23 (14.1%) | 1.0 |
| HIV | 7(3.0 %) | 3 (1.8%) | 0.85 |
| **Baseline ACH** |  |  |  |
| ACH mean | 2.36 ± 0.71 | 2.29 ± 0.73 | 0.3 |
| Mean number of sites<br>evaluated | 21.9 ± 15.6 | 19.4 ± 13.9 | 0.1 |
| **Change in ACH** |  |  |  |
| Months between xrays | 19.8 ± 5.9 | 17.6 ± 5.0 | <0.0001 |
| Mean change in ACH (mm) | -0.066 ± 0.393 | 0.034 ± 0.328 | 0.008 |
| Percent change in ACH | -1.74 ± 16.5% | 2.46 ± 14.6% | 0.01 |
| Annual percent change in ACH | -0.94 ± 12.5% | 1.33 ± 11.9% | 0.07 |

The duration between bitewings was greater in the pre-pandemic than post-pandemic group (19.8 + 5.9 vs 17.6 + 5.0 months, p<0.0001). The pre-pandemic group had a mean percentage loss of ACH of - 1.74 + 16.5 %, representing a gain in alveolar bone. In contrast, the post-pandemic group had a gain in ACH of 2.46 + 14.6 %, representing a loss in alveolar bone. We explored age, sex, race and ethnicity as covariates and found none of the to be associated with change in ACH. There remained a trend for greater annualized percent change in ACH in the post-pandemic vs pre-pandemic group (1.33 + 11.9% vs -0.94 + 12.5%, p=0.07), after accounting for differences in duration between xrays (Table 1) Among the 71 participants with 2 xrays in each time period, there were no statistically significant differences in the post- and pre-pandemic change in ACH (p=0.14) (Table 2). Similarly, there were no statistically significant differences in the annualized percent change in post- and pre-pandemic ACH (p=0.12).

**Table 2.** Change in ACH among participants (n=71) with two evaluable bitewing xrays.

|  | Mean Difference | 95% Confidence<br>Interval of mean | P value |
| --- | --- | --- | --- |
| **Pre-pandemic change in ACH (mm)<sup>1</sup>** | -0.07 ± 0.356 | -0.153, 0.013 | 0.1 |
| **Post-pandemic change in ACH (mm)<sup>2</sup>** | 0.015 ± 0.272 | -0.048, 0.078 | 0.64 |
| **Difference between post- and pre-pandemic change in ACH (mm)** | 0.082 ± 0.467 | -0.027, 0.192 | 0.14 |
| **Difference between post- and pre-pandemic change in ACH per year (mm)** | 0.057 ± 0.331 | -0.02, 0.135 | 0.15 |

<sup>1</sup> This is the difference in pre-pandemic mean ACH between exam 1 and exam 2
<sup>2</sup> This is the difference in pre-pandemic sum ACH between exam 3 and exam 4

## IV. Discussion

The deep learning accuracy results, when compared to dental professionals, highlight the immense potential of the deep learning application as a tool for precisely assessing severe versus non-severe ACHs both in clinical practice and for research purposes. It surpasses the capabilities of dental professionals in this specific diagnostic task. Furthermore, our data suggests that utilizing the DL-application could significantly save time for dental professionals when calculating and charting ACHs from xrays. A high percentage of dental professionals believe that the AI algorithm would be beneficial in their practice and believe that our application is accurate in calculating ACHs. There are a handful of commercially available AI integrated software packages that can calculate ACH, but based on our survey, very few dental professionals reported current use in their practice. This presents a great opportunity for the widespread use of deep learning algorithms in dental practices.

In this retrospective COVID study of data from a single institution, change in ACH was examined in a group of dental patients with paired bitewing xrays taken before and after the first cases of COVID were diagnosed in New York City. Radiographs examined in the post-pandemic group showed a greater alveolar bone loss compared to the pre-pandemic group. After accounting for between-group differences in duration time between bitewing xrays, a trend towards greater annualized percent change in the post-pandemic versus pre-pandemic group remained. However, the magnitude of change in ACH was modest (<0.1 mm), suggesting the lack of a clinically relevant impact of the COVID infection on alveolar bone loss. In our previous study, we found that patients who were diagnosed with COVID presented with increased ACH compared to controls that did not have a COVID positive test. In this current analysis we present longitudinal data comparing change in ACH during the pre- vs post-pandemic period.

Currently, we are not aware of other studies that have looked at the effect COVID may have on worsening alveolar bone loss. However, there have been several studies that have shown that periodontal disease affects the likelihood of becoming infected with COVID and effecting the severity of the infection [21, 22] It has been reported that patients with periodontitis had a 1.54 times higher risk of COVID complications and patients who had comorbidities were 2.49 times more likely to have COVID complications [21]. Furthermore, periodontal disease has been linked to COVID when looking at hospitalized COVID patients [22].

Various medical fields have benefited from deep learning models able to identify anatomical structures and detect pathological findings in radiographic images [1–3, 23]. Deep learning-based computer-aided diagnosis (CAD) for oral imaging has recently undergone significant development, but its implementation has been limited [3, 23, 24]. Some studies have attempted to measure the alveolar bone level by analyzing panoramic radiographs with deep learning models [3, 5–7]. Panoramic images provide a quick overview of the dentition, but they suffer from considerable distortion and a lack of detail, making accurate and precise diagnosis of periodontitis and other oral diseases difficult [3, 25, 26]. The current standard practice of visually evaluating intra-oral radiographs, is prone to errors . Advanced deep learning models on bitewing radiographs can provide an objective and dependable method for periodontal diagnosis. Lee, et al. utilized a deep learning semantic segmentation method to gauge the alveolar bone level on bitewing radiographs [3]. Our team has developed and implemented an advanced deep learning model that not only employs semantic segmentation neural networks but also object detection networks to precisely identify the ABCLs and cemento-enamel junctions (CEJs) for periodontal disease detection. The model was trained and validated using datasets curated by an Oral Radiologist (SM) from Columbia University College of Dental Medicine to set a gold standard for alveolar crest -height measurements.

There were several limitations in our study that could be addressed in future investigations. We were unable to compare xrays from patients with confirmed COVID to patients without COVID since most people had diagnostic testing performed outside of the hospital setting. If we limited our sample to patients with confirmed COVID tests within the hospital database, patients with positive tests would be biased for co-morbidities since only patients with severe COVID were hospitalized. For the purposes of this analysis, we assumed that everyone had at least one COVID infection during the post-pandemic period. The direction of bias is towards the null if fewer patients had COVID during this period. Additionally, our sample size was modest and we did not have complete data on comorbidities affecting periodontal health. For several patients in our sample, field of view differences between bitewings skewed the total ACH measurement values and therefore created false conclusions of increased bone loss over time. In order to improve the accuracy of future results, measuring ACH levels for specific teeth that match across every bitewing would eliminate any issues related to field of view differences between the bitewing images. Lastly, alveolar bone loss is a late manifestation of periodontal disease and is a chronic and dynamic process that can take several months to years to occur. A limitation to our study is that the window of time between bitewing images was limited. Further study in the future utilizing larger time windows between patient bitewings will demonstrate a more accurate representation of changes in the rate of alveolar bone loss.

Our analysis limited by a modest sample size and the inability to confirm COVID infection and timing of infection in our participants. While the “post-pandemic” group evidenced alveolar bone loss but the magnitude of change in ACH was modest (<0.1 mm), and provide assurance that COVID infection does not appear to have a large clinical impact on alveolar bone loss. Overall, our study demonstrates the successful training and validation of a deep learning program for ACH measurement based on a gold-standard training set read by an oral radiologist, as well as its utility and acceptability among dental professionals for both clinical and research purpose.

## Data Availability

All data produced in the present study are available upon reasonable request to the authors.

## V. Acknowledgments

**Funding**: This study was supported by the Columbia Biomedical Engineering Technology Accelerator (BiomedX) Program.

**Competing interests:** The authors declare that they have no competing interests.

**Data and materials availability:** Additional data related to this paper may be requested from the authors.

## References

[1] A. Esteva, A. Robicquet, B. Ramsundar, V. Kuleshov, M. DePristo, K. Chou, C. Cui, G. Corrado, S. Thrun, J. Dean, A guide to deep learning in healthcare, Nature medicine 25(1) (2019) 24–29.

[2] M.L. Giger, Machine learning in medical imaging, Journal of the American College of Radiology 15(3) (2018) 512–520.

[3] C.-T. Lee, T. Kabir, J. Nelson, S. Sheng, H.-W. Meng, T.E. Van Dyke, M.F. Walji, X. Jiang, S. Shams, Use of the Deep Learning Approach to Measure Alveolar Bone Level, arXiv preprint arXiv:2109.12115 (2021).

[4] O. Periodontitis, American Academy of Periodontology Task Force report on the update to the 1999 classification of periodontal diseases and conditions, J Periodontol 86(7) (2015) 835–838.

[5] J. Kim, H.-S. Lee, I.-S. Song, K.-H. Jung, DeNTNet: Deep Neural Transfer Network for the detection of periodontal bone loss using panoramic dental radiographs, Scientific reports 9(1) (2019) 17615.

[6] J. Krois, T. Ekert, L. Meinhold, T. Golla, B. Kharbot, A. Wittemeier, C. Dörfer, F. Schwendicke, Deep learning for the radiographic detection of periodontal bone loss, Scientific reports 9(1) (2019) 1–6.

[7] J.-H. Lee, D.-h. Kim, S.-N. Jeong, S.-H. Choi, Diagnosis and prediction of periodontally compromised teeth using a deep learning-based convolutional neural network algorithm, Journal of periodontal & implant science 48(2) (2018) 114–123.

[8] G. Mainas, L. Nibali, M. Ide, W.A. Mahmeed, K. Al-Rasadi, K. Al-Alawi, M. Banach, Y. Banerjee, A. Ceriello, M. Cesur, Associations between periodontitis, COVID-19, and cardiometabolic complications: molecular mechanisms and clinical evidence, Metabolites 13(1) (2022) 40.

[9] Z. Zheng, F. Peng, B. Xu, J. Zhao, H. Liu, J. Peng, Q. Li, C. Jiang, Y. Zhou, S. Liu, Risk factors of critical & mortal COVID-19 cases: A systematic literature review and meta-analysis, Journal of infection 81(2) (2020) e16–e25.

[10] E.S. Hosseini, N.R. Kashani, H. Nikzad, J. Azadbakht, H.H. Bafrani, H.H. Kashani, The novel coronavirus Disease-2019 (COVID-19): Mechanism of action, detection and recent therapeutic strategies, Virology 551 (2020) 1–9.

[11] H. Ejaz, A. Alsrhani, A. Zafar, H. Javed, K. Junaid, A.E. Abdalla, K.O. Abosalif, Z. Ahmed, S. Younas, COVID-19 and comorbidities: Deleterious impact on infected patients, Journal of infection and public health 13(12) (2020) 1833–1839.

[12] K. Sukumar, A. Tadepalli, Nexus between COVID-19 and periodontal disease, Journal of International Medical Research 49(3) (2021) 03000605211002695.

[13] V. Sampson, N. Kamona, A. Sampson, Could there be a link between oral hygiene and the severity of SARS-CoV-2 infections?, British dental journal 228(12) (2020) 971–975.

[14] L. Basso, D. Chacun, K. Sy, B. Grosgogeat, K. Gritsch, Periodontal diseases and COVID-19: a scoping review, European journal of dentistry 15(04) (2021) 768–775.

[15] B. Fernandes Matuck, M. Dolhnikoff, G.V. Maia, D. Isaac Sendyk, A. Zarpellon, S. Costa Gomes, A.N. Duarte-Neto, J.R. Rebello Pinho, M.S. Gomes-Gouvêa, S.C. Sousa, Periodontal tissues are targets for Sars-Cov-2: a post-mortem study, Journal of oral microbiology 13(1) (2021) 1848135.

[16] L. Vaughan, D. Veruttipong, J.G. Shaw, N. Levy, L. Edwards, M. Winget, Relationship of socio-demographics, comorbidities, symptoms and healthcare access with early COVID-19 presentation and disease severity, BMC infectious diseases 21 (2021) 1–10.

[17] G. Hajishengallis, Interconnection of periodontal disease and comorbidities: Evidence, mechanisms, and implications, Periodontology 2000 89(1) (2022) 9–18.

[18] C. Szegedy, S. Ioffe, V. Vanhoucke, A. Alemi, Inception-v4, inception-resnet and the impact of residual connections on learning, Proceedings of the AAAI conference on artificial intelligence, 2017.

[19] S. Ren, K. He, R. Girshick, J. Sun, Faster r-cnn: Towards real-time object detection with region proposal networks, Advances in neural information processing systems 28 (2015) 91–99.

[20] L.-C. Chen, G. Papandreou, I. Kokkinos, K. Murphy, A.L. Yuille, Deeplab: Semantic image segmentation with deep convolutional nets, atrous convolution, and fully connected crfs, IEEE transactions on pattern analysis and machine intelligence 40(4) (2017) 834–848.

[21] P. Koppolu, E.M. Genady, L.M. Albdeirat, F.A. Sebai, D.M. Alrashdi, A.S. Lingam, F.A.R. Alsada, F.I. Al-Khalifa, R.K. Abdelrahim, Association between severity of COVID-19, Periodontal health and disease in Riyadh subpopulation, The International Journal of Mycobacteriology 12(1) (2023) 33–37.

[22] Y. Wang, H. Deng, Y. Pan, L. Jin, R. Hu, Y. Lu, W. Deng, W. Sun, C. Chen, X. Shen, Periodontal disease increases the host susceptibility to COVID-19 and its severity: a Mendelian randomization study, Journal of translational medicine 19 (2021) 1–9.

[23] S. Shams, R. Platania, J. Zhang, J. Kim, K. Lee, S.-J. Park, Deep generative breast cancer screening and diagnosis, Medical Image Computing and Computer Assisted Intervention–MICCAI 2018: 21st International Conference, Granada, Spain, September 16-20, 2018, Proceedings, Part II 11, Springer, 2018, pp. 859–867.

[24] F. Schwendicke, T. Golla, M. Dreher, J. Krois, Convolutional neural networks for dental image diagnostics: A scoping review, Journal of dentistry 91 (2019) 103226.

[25] L. Åkesson, J. Håkansson, M. Rohlin, Comparison of panoramic and intraoral radiography and pocket probing for the measurement of the marginal bone level, Journal of clinical periodontology 19(5) (1992) 326–332.

[26] K. Hellén-Halme, A. Lith, X.-Q. Shi, Reliability of marginal bone level measurements on digital panoramic and digital intraoral radiographs, Oral radiology 36 (2020) 135–140.

